# Deep Learning to Estimate Cardiac Magnetic Resonance-Derived Left Ventricular Mass

**DOI:** 10.1101/2020.12.18.20248364

**Authors:** Shaan Khurshid, Samuel Friedman, James P. Pirruccello, Paolo Di Achille, Nathaniel Diamant, Christopher D. Anderson, Patrick T. Ellinor, Puneet Batra, Jennifer E. Ho, Anthony Philippakis, Steven A. Lubitz

## Abstract

**Background:** Cardiac magnetic resonance (CMR) is the gold standard for left ventricular hypertrophy (LVH) diagnosis. CMR-derived LV mass can be estimated using proprietary algorithms (e.g., inlineVF), but their accuracy and availability may be limited.

**Objective:** To develop an open-source deep learning model to estimate CMR-derived LV mass.

**Methods:** Within participants of the UK Biobank prospective cohort undergoing CMR, we trained two convolutional neural networks to estimate LV mass. The first (ML4H_reg_) performed regression informed by manually labeled LV mass (available in 5,065 individuals), while the second (ML4H_seg_) performed LV segmentation informed by inlineVF contours. We compared ML4H_reg_, ML4H_seg_, and inlineVF against manually labeled LV mass within an independent holdout set using Pearson correlation and mean absolute error (MAE). We assessed associations between CMR-derived LVH and prevalent cardiovascular disease using logistic regression adjusted for age and sex.

**Results:** We generated CMR-derived LV mass estimates within 38,574 individuals. Among 891 individuals in the holdout set, ML4H_seg_ reproduced manually labeled LV mass more accurately (r=0.864, 95% CI 0.847-0.880; MAE 10.41g, 95% CI 9.82-10.99) than ML4H_reg_ (r=0.843, 95% CI 0.823-0.861; MAE 10.51, 95% CI 9.86-11.15, p=0.01) and inlineVF (r=0.795, 95% CI 0.770-0.818; MAE 14.30, 95% CI 13.46-11.01, p<0.01). LVH defined using ML4H_seg_ demonstrated the strongest associations with hypertension (odds ratio 2.76, 95% CI 2.51-3.04), atrial fibrillation (1.75, 95% CI 1.37-2.20), and heart failure (4.53, 95% CI 3.16-6.33).

**Conclusions:** ML4H_seg_ is an open-source deep learning model providing automated quantification of CMR-derived LV mass. Deep learning models characterizing cardiac structure may facilitate broad cardiovascular discovery.

## Introduction

Left ventricular hypertrophy (LVH) is defined as pathologically increased LV mass^1^ and is consistently associated with increased risks of adverse cardiovascular events including heart failure,^1–3^ stroke,^1^ atrial fibrillation (AF),^4^ and sudden cardiac death.^5^ The gold standard for LVH diagnosis is cardiac magnetic resonance (CMR) imaging, which provides accurate and reproducible quantification of cardiac structure.^6^ However, traditional LV mass estimation using CMR requires LV segmentation, which is typically performed manually and requires substantial time and expertise.

The United Kingdom (UK) Biobank is a prospective cohort study comprised of over 500,000 individuals designed to facilitate broad-ranging research of diseases affecting middle-aged and older adults. Roughly 40,000 individuals have undergone prospective CMR acquisition, with additional imaging expected in another 60,000 individuals in the near future. However, manually quantified LV mass is available only within roughly 5,000 images,^7^ and additional measurements would be challenging to obtain at scale. Although automated quantification based on proprietary segmentation methods such as inlineVF (Siemens Healthineers, Erlangen, Germany) are accessible, previous work has suggested limited accuracy of resultant LV mass estimates, and the most recent software versions are not available to the majority of researchers.^8^ Therefore, a freely available method to facilitate accurate and automated quantification of LV mass using raw CMR images could enable impactful cardiovascular discovery research. In particular, deep learning methods may be well-suited for estimation of LV mass using CMR.

In the current study, we aimed to develop an open-source deep learning model to perform LV mass estimation from CMR images. We compared two separate approaches to LV mass estimation: 1) direct estimation trained on manually labeled LV mass values (Machine Learning for Health-Regression [ML4H_reg_]), and 2) identification of LV myocardial pixels followed by integration to obtain LV myocardial volume with subsequent conversion to LV mass (Machine Learning for Health-Segmentation [ML4H_seg_]) (**Figure 1**). We then compared the accuracy of both deep learning approaches to LV mass obtained using inlineVF within an independent holdout set using manually labeled LV mass as the gold standard.

**Figure 1.**
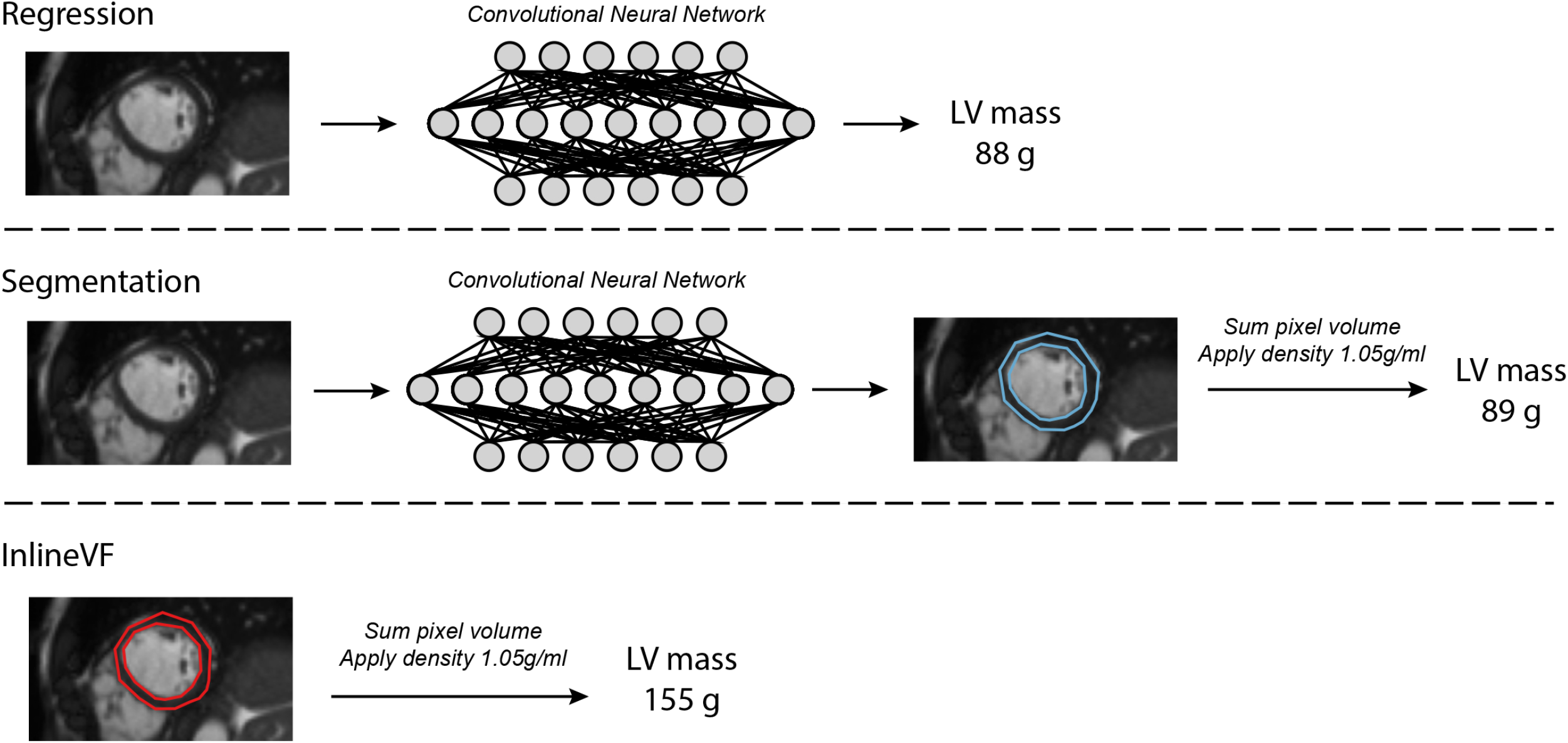
Overview of left ventricular mass algorithms Depicted is an overview of the three approaches to CMR-derived LV mass estimation compared in the current study. The top model utilizes deep learning-based regression trained by manually labeled LV mass. The middle model performs deep learning-based segmentation informed by inlineVF contours. The bottom model utilizes the inlineVF automated contours alone. For the deep learning segmentation and inlineVF models, LV segmentations were converted to LV mass by summing pixel volume and multiplying the density of LV myocardium (1.05g/mL, see text).

## Materials and Methods

### Study population

The UK Biobank is a population-based prospective cohort of 502,629 participants recruited between 2006-2010 in the United Kingdom primarily established to investigate the genetic and lifestyle determinants of disease. The design of the cohort has been described previously.^9,10^ Briefly, approximately 9.2 million individuals aged 40-69 living within 25 miles of the 22 assessment centers in England, Wales, and Scotland were invited, and 5.4% participated in the baseline assessment. Extensive questionnaire data, physical measures, and biological samples were collected at recruitment, with ongoing enhanced data collection in large subsets of the cohort, including repeated assessments and multimodal imaging. All participants are followed up for health outcomes through linkage to national health-related datasets. Participants provided written informed consent. The UK Biobank was approved by the UK Biobank Research Ethics Committee (reference number 11/NW/0382). Use of UK Biobank data (application 7089) was approved by the local Mass General Brigham Institutional Review Board.

### Cardiac magnetic resonance acquisition

For all analyses, we included individuals who underwent CMR during the UK Biobank imaging assessment and whose bulk CMR data were available for download as of 4-31-2019. The full CMR protocol of the UK Biobank has been described in detail previously.^11^ Briefly, all CMR examinations were performed in the United Kingdom on a clinical wide-bore 1.5 Tesla scanner (MAGNETOM Aera, Syngo Platform VD13A, Siemens Healthineers, Erlangen, Germany). All acquisitions used balanced steady-state free precession with typical parameters.

The contours extracted from the inlineVF algorithm and stored in each DICOM file’s metadata were further processed into pixel masks that labeled myocardium, LV cavity and background. The DICOM metadata stores the contour as polygons, which we processed into a pixel mask using the morphological image operators. The short axis CMR sequence in the UK Biobank contained between 6-13 short axis slices extending from base to apex. Height and width of the slices varied by individual but never exceeded 256 in either dimension. All CMR images were zero-padded to be three-dimensional tensors with shape (256, 256, 13). To facilitate the cross-entropy loss computation the three anatomical labels were one-hot encoded to be label masks with shape (256, 256, 13, 3). Each CMR image was normalized on a per-image basis to have mean zero and standard deviation one.

### Left ventricular mass models

We assessed two independent deep learning-based approaches to LV mass estimation. The first model was a 3D convolutional neural network regressor *ML4H*_*reg*_ trained with the manually annotated LV mass estimates provided by Petersen et al,^7^ *P(i)* to optimize the log cosh loss function, which behaves like L2 loss for small values and L1 loss for larger values:

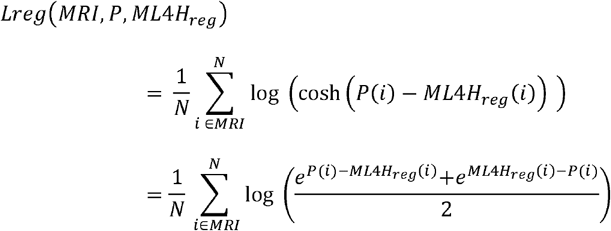

Here batch size, *N*, is 4 random samples from the training set of 3,178 after excluding testing and validation samples from the total 5,065 CMR images with LV mass values included in *P*. The second model *ML4H*_*seg*_ is a 3D semantic segmenter trained with the inlineVF contours to minimize *Lseg*, the per-pixel cross-entropy between the label and the model’s prediction.

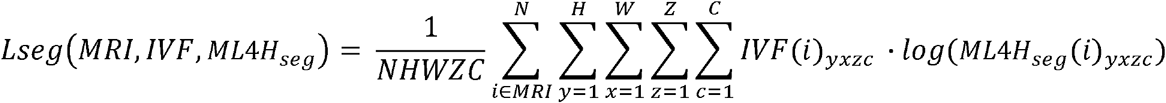

Here the batch size, *N*, was 4 from the total set of 33,071. Height, *H*, and width, *W*, are 256 voxels and there was a maximum of 13 *Z* slices along the short axis. There is a channel for each of the three labels, which were one-hot encoded in the training data, inlineVF (*IVF)*, and probabilistic values from the softmax layer of *ML4H*_seg_. Segmentation architectures used U-Net style long-range connections between early convolutional layers and deeper layers. Such an approach allows the final segmentation to use high-resolution local information with more abstract contextual features, both of which are critical for semantic segmentation. Since not all CMR images used the same pixel dimensions, we built models to incorporate pixel size values with their fully connected layers before making predictions. An overview of the architectures of both deep learning models is shown in **Supplemental Figure 1**.

For ML4H_reg_, an LV mass estimate was produced directly. To compute an LV mass estimate from ML4H_seg_ and inlineVF, we calculated LV mass based on the pixels predicted to be myocardium. Specifically, we multiplied the number of pixels corresponding to myocardium by the pixel depth (calculated to be 10mm using image metadata and visual confirmation, see **Supplemental Figure 2**) to yield total LV myocardial volume in milliliters. We then multiplied the predicted myocardial volume by the tissue density of LV myocardium, which is 1.05 g/cm^3^, to yield an estimate of LV mass.^12^ Given evidence of systematic overestimation in raw estimates obtained using both inlineVF and ML4H_seg_ (**Supplemental Figure 3**), we centered initial LV mass estimates using the observed mean LV mass of the manually labeled data within strata of sex.

All models were optimized using the ADAM variant of stochastic gradient descent with initial learning rate 1×10^−3^, exponential learning rate decay and batch size of 4 on K80 graphical processing units. Additional details regarding model training and evaluation are described in the **Supplemental Methods**. All models were implemented in tensorflow version 2.1.0 using the ML4H modeling framework.^13^ Model architectures, trained weights and more metrics are available at: https://github.com/broadinstitute/ml4h/tree/master/model_zoo/cardiac_mri_derived_left_ventricular_mass/.

### Disease associations

Given established associations between increased LV mass and the presence of LVH with cardiovascular disease, we assessed for associations between CMR-derived LV mass (using each method) and prevalent hypertension, atrial fibrillation, and heart failure. For these analyses, LVH was defined as LV mass index (LVMI) >72g/m^2^ in men and >55 g/m^2^ in women,^7^ and alternatively as the sex-specific 90^th^ percentile of LV mass.^1^ Indexing for body surface area was performed using the DuBois formula.^14^ Diseases were defined using self-report and inpatient ICD-9/10 codes (updated through 2020-03-31, **Supplemental Table 1**).

### Statistical analysis

The primary measure of LV mass estimation accuracy was the Pearson correlation between model-estimated LV mass values and hand-labeled LV mass within a holdout set independent of model training. We also calculated the mean absolute error (MAE) and analyzed agreement using Bland-Altman plots^15^ as secondary measures. Correlation coefficients were compared using Dunn and Clark’s *z* statistic^16^ for overlapping dependent correlations. Confidence intervals for MAE were obtained using 1,000-iteration bootstrapping. A linear recalibration formula to correct for bias in inlineVF-based measurements was obtained by regressing manually labeled LV mass on LV mass estimates obtained using inlineVF. Associations between LV mass and LVH with prevalent disease were assessed using logistic regression models with adjustment for age and sex.

To assess the behavior of the deep learning segmentation model, we generated saliency maps (maps denoting CMR regions identified as myocardium). Statistical analyses were performed using R v3.5 (packages ‘data.table’, ‘ggplot2’, ‘epiR’, ‘pROC’, ‘nricens’).^17,18^ All two-tailed p-values <0.05 were considered statistically significant.

## Results

Within 33,071 individuals who underwent CMR, we trained models to derive CMR-based LV mass using deep learning regression (ML4H_reg_) and segmentation (ML4H_seg_). The mean age was 64 ± 8 years and 52% were female. Other baseline characteristics of the training and test sets are shown in **Table 1**.

**Table 1.** Baseline characteristics

|  | **Training set**<br>**(N=33,071)** | **Holdout set**<br>**(N=5,393)** |
| --- | --- | --- |
| Age | 64.2 ± 7.5 | 63.6 ± 7.7 |
| Female | 17,183 (52.0%) | 2,847 (52.8%) |
| Race/Ethnicity | - | - |
| White | 32,013 (96.8%) | 5,235 (97.1%) |
| Asian or Pacific Islander | 446 (1.3%) | 61 (1.1%) |
| Black | 207 (0.6%) | 29 (0.5%) |
| Mixed | 151 (0.5%) | 23 (0.4%) |
| Other | 159 (0.5%) | 27 (0.5%) |
| Unknown | 95 (0.3%) | 18 (0.3%) |
| Systolic blood pressure (mmHg) | 135 ± 18 | 135 ± 18 |
| Diastolic blood pressure (mmHg) | 81 ± 10 | 81 ± 10 |
| HTN | 10,122 (30.6%) | 1,572 (29.1%) |
| Diabetes | 1288 (3.9%) | 186 (3.4%) |
| Heart failure | 191 (0.6%) | 26 (0.5%) |
| Myocardial infarction | 686 (2.1%) | 101 (1.9%) |
| CMR-derived LV mass (inlineVF, g) | 154.9 ± 38.5 | 154.9 ± 38.2 |
| CMR-derived LV mass (inlineVF centered, g) | 90.0 ± 33.1 | 90.1 ± 32.8 |
| CMR-derived LV mass (regression, g) | 88.2 ± 16.2 | 87.6 ± 15.6 |
| CMR-derived LV mass (segmentation, g) | 88.9 ± 28.4 | 89.1 ± 27.8 |

Initial LV mass estimates obtained using inlineVF and ML4H_seg_ demonstrated evidence of systematic overestimation, which was corrected after centering each distribution upon the mean observed manually labeled LV mass. The distributions of LV mass stratified by sex using each method are shown in **Figure 2**. In an independent holdout set of 891 individuals with manually labeled LV mass estimates available, ML4H_seg_ had favorable correlation with manually labeled LV mass (r=0.864, 95% CI 0.847-0.880; MAE 10.41g, 95% CI 9.82-10.99) as compared to ML4H_reg_ (r=0.843, 95% CI 0.823-0.861; MAE 10.51, 95% CI 9.86-11.15, p=0.01) and centered inlineVF (r=0.795, 95% CI 0.770-0.818; MAE 14.30, 95% CI 13.46-11.01, p<0.01, **Figure 3**). Bland-Altman plots demonstrated reasonable agreement between both ML4H_seg_ and ML4H_reg_ and manually labeled LV mass, although ML4H_reg_ tended to progressively underestimate greater LV mass values (**Figure 4**). Saliency maps suggested that the ML4H_seg_ appropriately identified areas of LV myocardium for LV mass estimation (**Supplemental Figure III** and **Supplemental Methods**). Correlations between manually labeled LV mass with inlineVF and the ML4H_seg_ prior to mean centering are shown in **Supplemental Figures IV-V**. Correlation between manually labeled LV mass and inlineVF additionally adjusted using linear recalibration was slightly improved (r=0.838, 95% CI 0.817-0.856) and is shown in **Supplemental Figure V**. A bias-corrected inlineVF LV mass can be calculated using the following equation: 0.543487 x *unadjusted inlinevF Lv mass* + 5.808005.

**Figure 2.**
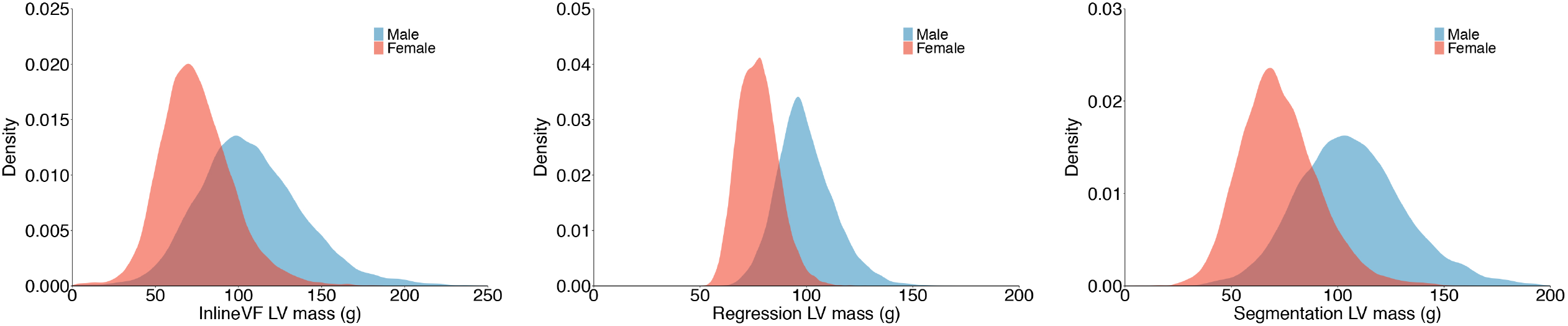
Distributions of CMR-derived LV mass obtained using each estimation method Depicted are density plots showing the distribution of CMR-derived LV mass (x-axis) using mean-centered inlineVF (left panel), the deep learning regression model (middle panel), and the deep learning segmentation model (right panel). Results are shown for the full sample with available CMR imaging (N=38,464).

**Figure 3.**
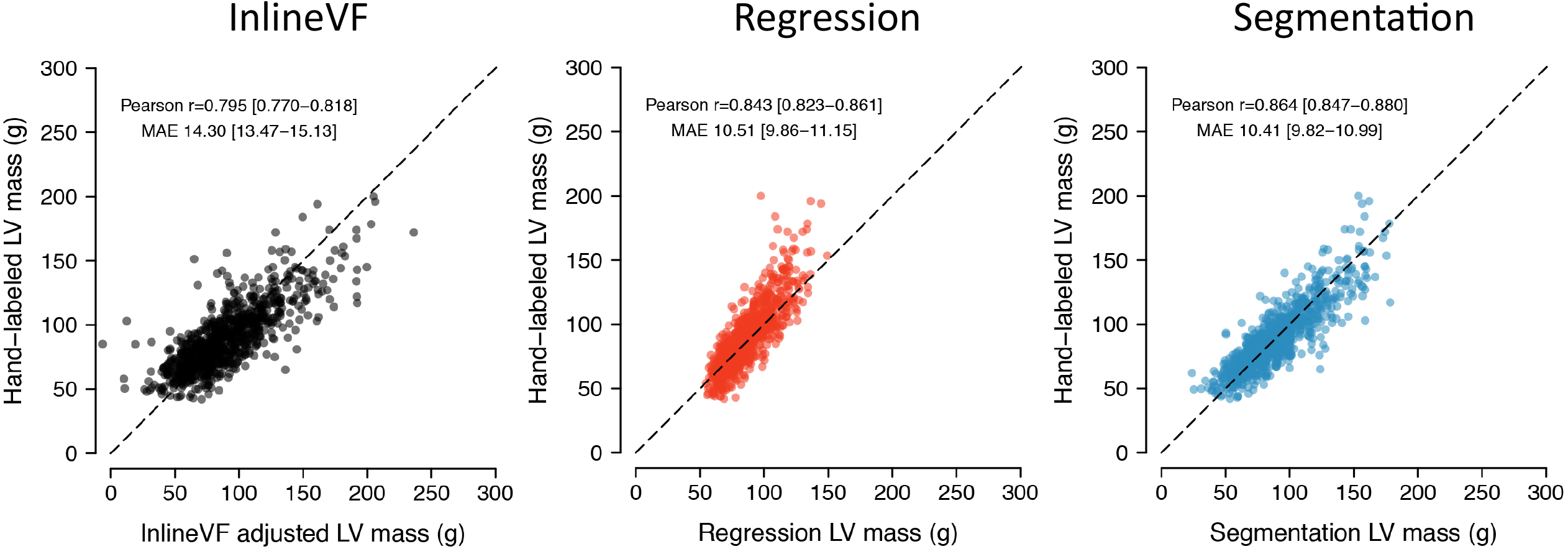
Correlation between manually labeled left ventricular mass and derived left ventricular mass estimated using each model Depicted are plots illustrating the correlation between manually labeled LV mass (y-axis) and CMR-derived LV mass using inlineVF (left panel), deep learning regression (middle panel), and deep learning segmentation (right panel). Results are shown among individuals within the test set independent of model training. Estimates for inlineVF and the segmentation model are displayed after centering the distribution upon the observed sex-stratified mean manually labeled LV mass (see text).

**Figure 4.**
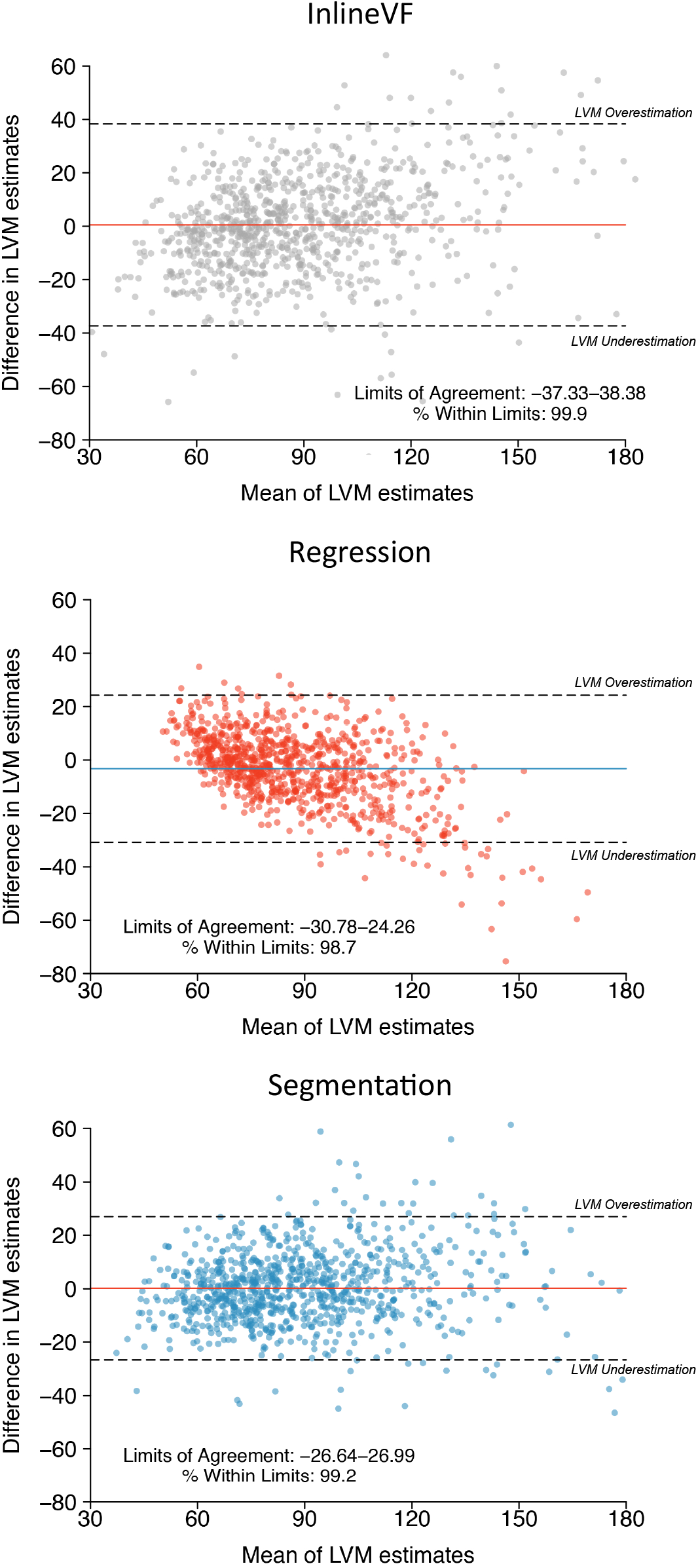
Bland-Altman plots comparing manually labeled left ventricular mass and derived left ventricular mass using each model Depicted are Bland-Altman plots^15^ showing agreement between manually labeled LV mass and LV mass estimated using inlineVF (top), ML4H_reg_ (middle), and ML4H_seg_ (bottom). Estimates using inlineVF and ML4H_seg_ are depicted after mean centering (see text). In each plot, each point represents a paired observation (i.e., the manually labeled LV mass estimate and the model predicted LV mass estimate). The x-axis depicts increasing mean of the paired observations. The y-axis depicts the difference between the paired observations, with negative values representing pairs in which manually labeled LV mass was larger than model predicted LV mass (underestimation using the model). The colored horizontal line shows the overall mean difference within each sample, and the hashed horizontal lines show the upper and lower bounds of the mean difference (defined as ±1.96 standard deviations [SD] of the difference). The corresponding bounds (a surrogate for level of agreement) and the proportion of observations within those bounds are depicted on each plot. A total of 13 (inlineVF), 1 (ML4H_reg_), and 1 (ML4H_seg_) outlying observations are not depicted for graphical purposes.

### Associations between CMR-based LV mass and prevalent disease

We assessed for associations between CMR-derived LV mass and prevalent cardiovascular disease. At the time of CMR acquisition, there were 11,271 prevalent hypertension, 1,053 atrial fibrillation, and 236 heart failure events. When compared to the other approaches, LVH defined using ML4H_seg_ consistently demonstrated the strongest associations with hypertension (odds ratio [OR] 2.76, 95% CI 2.51-3.04), atrial fibrillation (1.75, 95% CI 1.37-2.20), and heart failure (4.53, 95% CI 3.16-6.33, **Table 2**).

**Table 2.** Associations between deep learning segmentation-derived LV mass index and prevalent disease

|  |  | Odds ratio with covariate (95% CI) |  |  |
| --- | --- | --- | --- | --- |
|  | N events* | LVMI (per 1 SD) | LVH | LVH (90<sup>th</sup> percentile) |
| *Hypertension* |  |  |  |  |
| InlineVF | 11,271 | 1.43 (1.39-1.47) | 2.30 (2.15-2.46) | 2.33 (2.17-2.50) |
| Regression | 11,271 | 1.27 (1.24-1.30) | 1.67 (1.38-2.01) | 1.64 (1.53-1.76) |
| Segmentation | 11,271 | 1.55 (1.51-1.59) | 2.76 (2.51-3.04) | 2.39 (2.23-2.57) |
| *Atrial fibrillation* |  |  |  |  |
| InlineVF | 1,053 | 0.99 (0.93-1.05) | 1.19 (0.99-1.44) | 1.27 (1.04-1.53) |
| Regression | 1,053 | 1.00 (0.93-1.07) | 1.13 (0.59-1.93) | 0.99 (0.80-1.21) |
| Segmentation | 1,053 | 1.13 (1.06-1.21) | 1.75 (1.37-2.20) | 1.61 (1.34-1.93) |
| *Heart failure* |  |  |  |  |
| InlineVF | 236 | 1.42 (1.26-1.60) | 2.82 (2.07-3.78) | 2.90 (2.12-3.90) |
| Regression | 236 | 1.37 (1.21-1.55) | 3.60 (1.50-7.27) | 2.31 (1.66-3.14) |
| Segmentation | 236 | 1.67 (1.49-1.90) | 4.53 (3.16-6.33) | 3.59 (2.66-4.80) |
| *Total N=37,261 with available phenotypic data and CMR-derived LV mass estimates obtained using each method |  |  |  |  |

## Discussion

Within over 30,000 individuals with CMR imaging performed as part of the UK Biobank prospective cohort study, we developed and tested two deep learning-based approaches to automated LV mass estimation. When compared to one other and the proprietary D13A inlineVF automated segmentation software, ML4H_seg_ demonstrated the greatest correlation (86%) and lowest estimation error (approximately 10g) when compared against manually labeled LV mass. Importantly, greater CMR-derived LV mass obtained using our segmentation model had the strongest associations with prevalent hypertension, atrial fibrillation, and heart failure. We have shared our model architecture publicly and aim to return CMR-derived LV mass values to the UK Biobank for use by other researchers in order to facilitate future cardiovascular discovery utilizing rich cardiac structural imaging features.

Our study supports and extends previous work demonstrating the potential for deep learning to provide automated quantification of imaging phenotypes. A study by Aung et al.^19^ utilized a combination of deep learning and manual segmentation to extend LV mass estimates to approximately 16,000 UK Biobank images, in order to facilitate genetic analyses. Similarly, Bai et al.^20^ utilized deep learning to estimate several cardiac structural features to enable broad phenotypic association testing. In contrast to previous work, we explicitly compared several approaches to automated LV mass estimation, observing that a deep learning segmentation model demonstrated favorable performance when compared to deep learning-based regression and a recalibrated inlineVF-based method. Furthermore, we validated the performance of our best performing algorithm by assessing correlation and agreement against manually labeled estimates as well as assessing for expected associations with prevalent disease.

Our results provide insight into the comparative accuracy of potential methods to estimate cardiac structural features, suggesting that automated segmentation may provide superior performance. Specifically, we compared three approaches to automated LV mass estimation: deep learning-based segmentation (ML4H_seg_), deep learning-based regression on hand-labeled LV mass estimates (ML4H_reg_), and use of the automated contours provided by the inlineVF D13A proprietary software. We observed favorable accuracy using ML4H_seg_, which more closely mirrors the manual process of clinical LV mass estimation, in which cardiac radiologists manually label pixels as LV myocardium. Nevertheless, our results demonstrate that a regression approach can achieve reasonable accuracy, which may be improved as more training data becomes available. Future work is needed to better understand the relative strengths and weaknesses of various approaches to deep learning-based cardiac structural characterization, as well as to assess the comparative generalizability of such approaches when transferred to external datasets.

The current work highlights the potential for deep learning to derive clinically relevant imaging phenotypes in an efficient and automated manner. Increased LV mass and LVH have long been implicated as important risk factors for adverse cardiovascular events.^1–5^ The detection of LVH is clinically relevant since the majority of cases are related to hypertension, for which treatment can lead to regression of hypertrophy and improvement in cardiovascular risk profile.^21,22^ The associations and effect sizes we observed between CMR-derived LVH and prevalent hypertension, atrial fibrillation and heart failure were consistently strongest using ML4H_seg_ and are broadly consistent with prior studies.^2,4^ On balance, our findings suggest that deep learning may facilitate recognition of clinically relevant degrees of increased LV mass in a manner deployable at scale.

We submit that our findings may directly enable future cardiovascular research focused on CMR-derived LV mass, and potentially additional imaging phenotypes. The UK Biobank has performed CMR in over 35,000 individuals, with imaging expected to extend to nearly 100,000 individuals in the near future. Although LV mass and LVH reflect clinically important aspects of cardiac structure, quantification of LV mass using gold-standard CMR is traditionally performed manually, which is time-consuming and requires specialized expertise. Although UK Biobank images include the inlineVF D13A automated contours which may be used to estimate LV mass, our findings support previous studies demonstrating substantial overestimation.^8^ Furthermore, inlineVF is a proprietary algorithm whose latest versions are not accessible to all investigators.^8^ To this end, our deep learning model ML4H_seg_ provides more accurate and substantially less biased estimates, and the code underlying the model is available for public use. For investigators opting not to deploy our deep learning model, we also provide a formula to obtain linearly adjusted LV mass using inlineVF D13A, which demonstrated considerable bias correction and only moderately lower correlation as compared to our segmentation model.

Our study should be interpreted in the context of design. First, although the correlation between our deep learning model and manually labeled LV mass was very good (86%), it was not perfect, which may result in some misclassification of LV mass. Nevertheless, it was the best performing model of the three approaches tested, both in terms of correlation, agreement, and absolute error. Second, the number of manually labeled LV mass values available to train and evaluate models was relatively limited. Future models trained on a greater number of ground truth examples may enable the development of more accurate deep learning-based LV mass estimates. Third, the distribution of LV mass in the UK Biobank is lower than that observed in other settings.^2,7^ As a result, external validation of ML4H_seg_ would be needed prior to deployment in datasets other than the UK Biobank. Fourth, the degree of improvement observed using our segmentation model as opposed to inlineVF is relatively modest, especially after linear correction of inlineVF-based estimates. However, we submit that a 3-point increase in correlation adds substantive value, and the linear correction we provide is based on manually labeled LV mass estimates from a relatively small proportion of the total images available in the UK Biobank. As a result, the performance of linearly corrected inlineVF may decrease when applied to the remaining images and future releases.

## Conclusions

Utilizing a unique resource of CMR images obtained within over 35,000 individuals, we developed ML4H_seg_ – a deep learning segmentation model that provides automated LV mass estimation with favorable accuracy as compared to deep learning regression or the inlineVF proprietary algorithm. Importantly, model-derived LV mass estimates demonstrated expected associations with cardiovascular disease. We have made our algorithm publicly available for future use, and submit that such deep learning approaches may facilitate broad cardiovascular discovery by enabling future analyses of CMR-derived cardiac structural phenotypes available at scale.

## Data Availability

Model architectures, trained weights and more metrics are available at: https://github.com/broadinstitute/ml4h/tree/master/model_zoo/cardiac_mri_derived_left_ventricular_mass/.

---

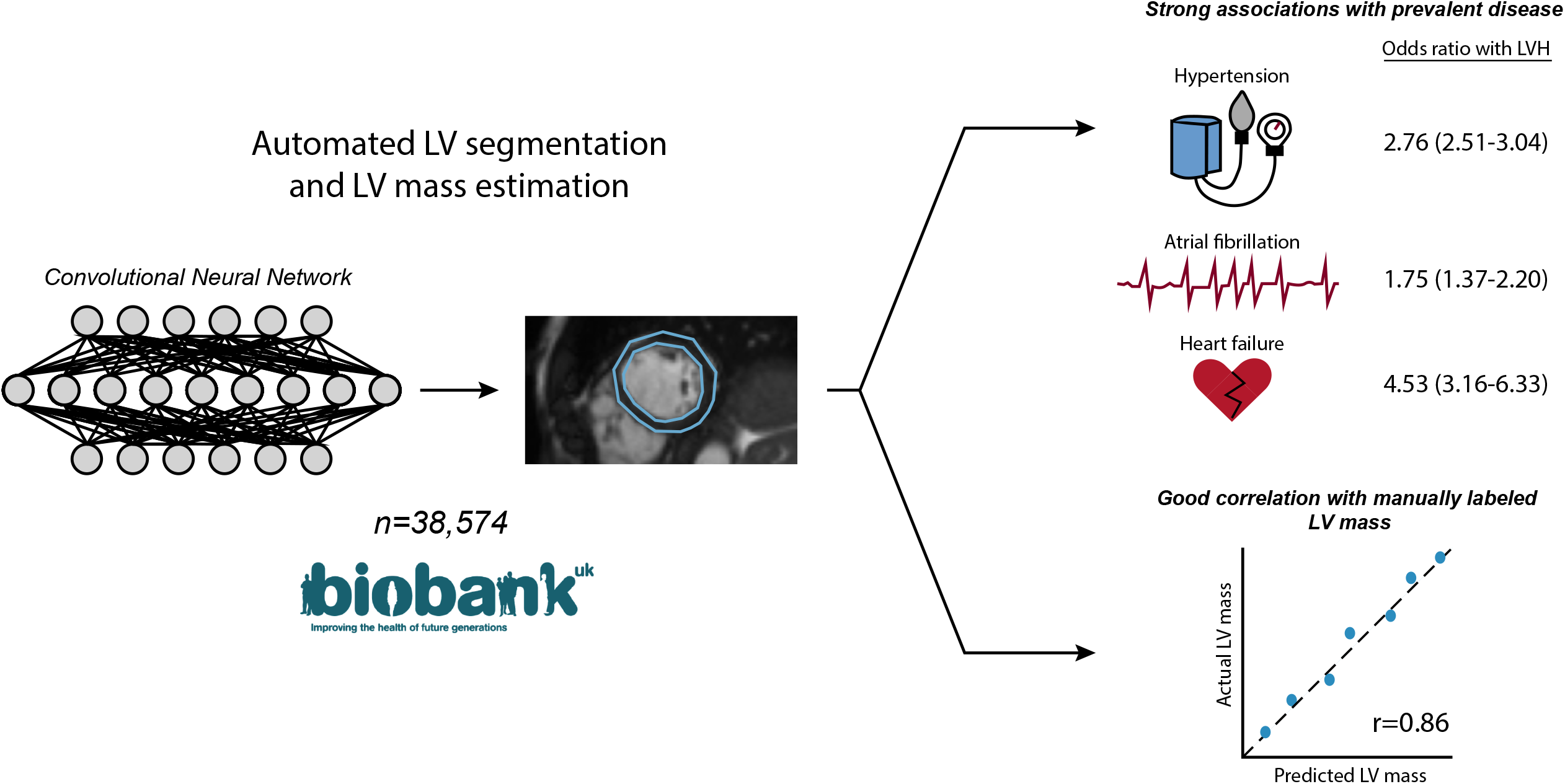

## References

1. Bluemke DA, Kronmal RA, Lima JAC, et al.: The relationship of left ventricular mass and geometry to incident cardiovascular events: the MESA (Multi-Ethnic Study of Atherosclerosis) study. J Am Coll Cardiol 2008; 52:2148–2155.

2. Kawel-Boehm N, Kronmal R, Eng J, et al.: Left Ventricular Mass at MRI and Long-term Risk of Cardiovascular Events: The Multi-Ethnic Study of Atherosclerosis (MESA). Radiology 2019; 293:107–114.

3. Lazzeroni D, Rimoldi O, Camici PG: From Left Ventricular Hypertrophy to Dysfunction and Failure. Circ J 2016; 80:555–564.

4. Chrispin J, Jain A, Soliman EZ, et al.: Association of electrocardiographic and imaging surrogates of left ventricular hypertrophy with incident atrial fibrillation: MESA (Multi-Ethnic Study of Atherosclerosis). J Am Coll Cardiol 2014; 63:2007– 2013.

5. Haider AW, Larson MG, Benjamin EJ, Levy D: Increased left ventricular mass and hypertrophy are associated with increased risk for sudden death. J Am Coll Cardiol 1998; 32:1454–1459.

6. Lenstrup M, Kjaergaard J, Petersen CL, Kjaer A, Hassager C: Evaluation of left ventricular mass measured by 3D echocardiography using magnetic resonance imaging as gold standard. Scand J Clin Lab Invest 2006; 66:647–657.

7. Petersen SE, Aung N, Sanghvi MM, et al.: Reference ranges for cardiac structure and function using cardiovascular magnetic resonance (CMR) in Caucasians from the UK Biobank population cohort. J Cardiovasc Magn Reson 2017; 19:18.

8. Suinesiaputra A, Sanghvi MM, Aung N, et al.: Fully-automated left ventricular mass and volume MRI analysis in the UK Biobank population cohort: evaluation of initial results. Int J Cardiovasc Imaging 2018; 34:281–291.

9. Sudlow C, Gallacher J, Allen N, et al.: UK biobank: an open access resource for identifying the causes of a wide range of complex diseases of middle and old age. PLoS Med 2015; 12:e1001779.

10. Littlejohns TJ, Sudlow C, Allen NE, Collins R: UK Biobank: opportunities for cardiovascular research. Eur Heart J 2019; 40:1158–1166.

11. Petersen SE, Matthews PM, Francis JM, et al.: UK Biobank’s cardiovascular magnetic resonance protocol. J Cardiovasc Magn Reson 2016; 18:8.

12. Myerson SG, Bellenger NG, Pennell DJ: Assessment of left ventricular mass by cardiovascular magnetic resonance. Hypertension 2002; 39:750–755.

13. ML4CVD Group: Machine Learning for Health (ML4H). https://github.com/broadinstitute/ml.GitHub 2020;.

14. Du Bois D, Du Bois EF: A formula to estimate the approximate surface area if height and weight be known. 1916. Nutrition 1989; 5:303–311; discussion 312-313.

15. Bland JM, Altman DG: Measuring agreement in method comparison studies. stat methods med res 1999; 8:135–160.

16. Dunn OJ, Clark V: Correlation Coefficients Measured on the Same Individuals. Journal of the American Statistical Association 1969; 64:366–377.

17. R Core Team (2015). R: A language and environment for statistical computing. R Foundation for Statistical Computing Vienna, Austria. URL https://www.R-project.org/.

18. Dowle M, Srinivasan A, Gorecki J, et al.: data.table: Extension of “data.frame”. Version 1.12.6. https://CRAN.R-project.org/package=data.table.

19. Aung N, Vargas JD, Yang C, et al.: Genome-Wide Analysis of Left Ventricular Image-Derived Phenotypes Identifies Fourteen Loci Associated With Cardiac Morphogenesis and Heart Failure Development. Circulation 2019; 140:1318–1330.

20. Bai W, Suzuki H, Huang J, et al.: A population-based phenome-wide association study of cardiac and aortic structure and function. Nat Med 2020; 26:1654–1662.

21. Dahlöf B, Pennert K, Hansson L: Reversal of Left Ventricular Hypertrophy in Hypertensive Patients. American Journal of Hypertension 1992; 5:95–110.

22. Okin PM, Devereux RB, Jern S, et al.: Regression of electrocardiographic left ventricular hypertrophy during antihypertensive treatment and the prediction of major cardiovascular events. JAMA 2004; 292:2343–2349.

