## Supplemental Material for "Deep Learning to Estimate Cardiac Magnetic Resonance-Derived Left Ventricular Mass"

**Index**

Table 1. Clinical factor definitions	1

**Figure 1.** Deep learning regression and segmentation model architectures	5

Figure 2. Determination of short-axis CMR slice thickness	6

**Figure 3.** Saliency map of left ventricular mass semantic segmentation model	7

**Figure 4.** Correlation between manually labeled left ventricular mass and estimated LV mass using deep learning segmentation prior to adjustment	8

**Figure 5.** Correlation between manually labeled left ventricular mass and estimated LV mass using inlineVF with and without adjustment	9

Methods. Deep learning model training and evaluation	10

Supplemental References	14

**Table 1**. Clinical factor definitions

| **Phenotype** | **Data fields** | **Field names** | **Data codes** | **Data code definitions** |
| --- | --- | --- | --- | --- |
| **UK Biobank** | | | | |
| Atrial fibrillation | 20002 | Non-cancer illness code, self-reported | 1471, 1483 | Atrial fibrillation, Atrial flutter |
| Atrial fibrillation | 20004 | Operation code, self-reported | 1524 | Cardioversion |
| Atrial fibrillation | 41202  41204 40001 40002 | Diagnoses - main ICD10  Diagnoses – secondary ICD10  Underlying (primary) cause of death: ICD10, Contributory (secondary) cause of death: ICD10 | I48, I48.0, I48.1, I48.2, I48.3, I48.4, I48.9 | Atrial fibrillation and flutter, Paroxysmal atrial fibrillation, Persistent atrial fibrillation, Chronic atrial fibrillation, Typical atrial flutter, Atypical atrial flutter, Atrial fibrillation and flutter, unspecified |
| Atrial fibrillation | 41203  41205 | Diagnosis - main ICD9  Diagnoses - secondary ICD9 | 4273 | Atrial fibrillation and flutter |
| Atrial fibrillation | 41200  41210 | Operative procedures – main OPCS  Operative procedures – secondary OPCS | K57.1, K62.1, K62.2, K62.3, K62.4, X50.1, X50.2 | Percutaneous transluminal ablation of atrioventricular node, Percutaneous transluminal ablation of pulmonary vein to left atrium conducting system, Percutaneous transluminal ablation of atrial wall for atrial flutter, Percutaneous transluminal ablation of conducting system of heart for atrial flutter NEC, Percutaneous transluminal internal cardioversion NEC, Direct current cardioversion, External cardioversion NEC |
| Diabetes | 2443 | Diabetes diagnosed by doctor | 1 | Yes |
| Diabetes | 20002 | Non-cancer illness code, self-reported | 1220, 1221, 1222, 1223 | Diabetes, Gestational diabetes, Type 1 diabetes, Type 2 diabetes |
| Diabetes | 2986 | Insulin use within one year | 1 | Started insulin within one year diagnosis of diabetes - Yes |
| Diabetes | 6177 | Medication for cholesterol, blood pressure or diabetes | 3 | Insulin |
| Diabetes | 6153 | Medication for cholesterol, blood pressure, diabetes, or take exogenous hormones | 3 | Insulin |
| Diabetes | 41202  41204 40001 40002 | Diagnoses - main ICD10  Diagnoses – secondary ICD10  Underlying (primary) cause of death: ICD10, Contributory (secondary) cause of death: ICD10 | E10, E10.0, E10.1, E10.2, E10.3, E10.4, E10.5, E10.6, E10.7, E10.8, E10.9, E11, E11.0, E11.1, E11.2, E11.3, E11.4, E11.5, E11.6, E11.7, E11.8, E11.9, E12, E12.1, E12.8, E12.9, E13, E13.1, E13.2, E13.3, E13.5, E13.6, E13.7, E13.8, E13.9, E14, E14.0, E14.1, E14.2, E14.3, E14.4, E14.5, E14.6, E14.7, E14.8, E14.9 | Insulin-dependent diabetes mellitus, Insulin-dependent diabetes mellitus with coma, Insulin-dependent diabetes mellitus with ketoacidosis, Insulin-dependent diabetes mellitus with renal complications, Insulin-dependent diabetes mellitus with ophthalmic complications, Insulin-dependent diabetes mellitus with neurological complications, Insulin-dependent diabetes mellitus with peripheral circulatory complications, Insulin-dependent diabetes mellitus with other specified complications, Insulin-dependent diabetes mellitus with multiple complications, Insulin-dependent diabetes mellitus with unspecified complications, Insulin-dependent diabetes mellitus without complications, Non-insulin-dependent diabetes mellitus, Non-insulin-dependent diabetes mellitus - with coma, Non-insulin-dependent diabetes mellitus - with ketoacidosis, Non-insulin-dependent diabetes mellitus - with renal complications, Non-insulin-dependent diabetes mellitus - with ophthalmic complications, Non-insulin-dependent diabetes mellitus - with neurological complications, Non-insulin-dependent diabetes mellitus - with peripheral circulatory complications, Non-insulin-dependent diabetes mellitus - with other specified complications, Non-insulin-dependent diabetes mellitus - with multiple complications, Non-insulin-dependent diabetes mellitus - with unspecified complications, Non-insulin-dependent diabetes mellitus - without complications, Malnutrition-related diabetes mellitus, Malnutrition-related diabetes mellitus with ketoacidosis, Malnutrition-related diabetes mellitus with unspecified complications, Malnutrition-related diabetes mellitus without complications, Other specified diabetes mellitus, Other specified diabetes mellitus with ketoacidosis, Other specified diabetes mellitus with renal complications, Other specified diabetes mellitus with ophthalmic complications, Other specified diabetes mellitus with peripheral circulatory complications, Other specified diabetes mellitus with other specified complications, Other specified diabetes mellitus with multiple complications, Other specified diabetes mellitus with unspecified complications, Other specified diabetes mellitus without complications, Unspecified diabetes mellitus, Unspecified diabetes mellitus with coma, Unspecified diabetes mellitus with ketoacidosis, Unspecified diabetes mellitus with renal complications, Unspecified diabetes mellitus with ophthalmic complications, Unspecified diabetes mellitus with neurological complications, Unspecified diabetes mellitus with peripheral circulatory complications, Unspecified diabetes mellitus with other specified complications, Unspecified diabetes mellitus with multiple complications, Unspecified diabetes mellitus with unspecified complications, Unspecified diabetes mellitus without complications |
| Diabetes | 41203  41205 | Diagnosis - main ICD9  Diagnoses - secondary ICD9 | 2500, 25000, 25001, 25009, 2501, 25011, 25019, 2503, 2504, 2505, 25099 | Diabetes mellitus without mention of complication, Diabetes mellitus without mention of complication (adult-onset type), Diabetes mellitus without mention of complication (juvenile type), Diabetes mellitus without mention of compl. (adult/juvenile unspec.), Diabetes with ketoacidosis, Diabetes with ketoacidosis (juvenile type), Diabetes with ketoacidosis (adult/juvenile unspec.), Diabetes with renal manifestations, Diabetes with ophthalmic manifestations, Diabetes with neurological manifestations, Diabetes with unspecified complications (unspecified onset) |
| Heart Failure | 20002 | Non-cancer illness code, self-reported | 1076, 1079, 1588 | Heart failure/pulmonary oedema, Cardiomyopathy, Hypertrophic cardiomyopathy (hcm / hocm) |
| Heart Failure | 41202  41204  40001  40002 | Diagnoses - main ICD10  Diagnoses – secondary ICD10  Underlying (primary) cause of death: ICD10, Contributory (secondary) cause of death: ICD10 | I11.0, I13.0, I13.2, I25.5, I42.0, I42.1, I42.2, I42.5, I42.8, I42.9, I50, I50.0, I50.1, I50.9 | Hypertensive heart disease with (congestive) heart failure, Hypertensive heart and renal disease with (congestive) heart failure, Hypertensive heart and renal disease with both (congestive) heart failure and renal failure, Hypertensive heart disease with (congestive) heart failure, Hypertensive heart and renal disease with (congestive) heart failure, Hypertensive heart and renal disease with both (congestive) heart failure and renal failure, Ischaemic cardiomyopathy, Dilated cardiomyopathy, Obstructive hypertrophic cardiomyopathy, Other hypertrophic cardiomyopathy, Other restrictive cardiomyopathy, Other cardiomyopathies, Cardiomyopathy, unspecified, Heart failure, Congestive heart failure, Left ventricular failure, Heart failure, unspecified |
| Hypertension | 20002 | Non-cancer illness code, self-reported | 1065, 1072 | Hypertension, essential hypertension |
| Hypertension | 6150 | Vascular/heart problems diagnosed by doctor | 4 | High blood pressure |
| Hypertension | 41202  41204 40001 40002 | Diagnoses - main ICD10  Diagnoses – secondary ICD10  Underlying (primary) cause of death: ICD10, Contributory (secondary) cause of death: ICD10 | I10, I11, I11.0, I11.9, I12, I12.0, I12.9, I13, I13.0, I13.1, I13.2, I13.9, I15, I15.0, I15.1, I15.2, I15.8, I15.9 | Essential (primary) hypertension, Hypertensive heart disease, Hypertensive heart disease with (congestive) heart failure, Hypertensive heart disease without (congestive) heart failure, Hypertensive renal disease, Hypertensive renal disease with renal failure, Hypertensive renal disease without renal failure, Hypertensive heart and renal disease, Hypertensive heart and renal disease with (congestive) heart failure, Hypertensive heart and renal disease with renal failure, Hypertensive heart and renal disease with both (congestive) heart failure and renal failure, Hypertensive heart and renal disease, unspecified, Secondary hypertension, Renovascular hypertension, Hypertension secondary to other renal disorders, Hypertension secondary to endocrine disorders, Other secondary hypertension, Secondary hypertension, unspecified |
| Hypertension | 41203  41205 | Diagnoses - main ICD9  Diagnoses – secondary ICD9 | 401, 4010, 4011, 4019, 402, 4020, 4021, 4029, 403, 4030, 4031, 4039, 404, 4040, 4041, 4049, 405, 4050, 4051, 4059 | Essential hypertension, Essential hypertension, specified as malignant, Essential hypertension, specified as benign, Essential hypertension, not specified as malignant or benign, Hypertensive heart disease, Hypertensive heart disease, specified as malignant, Hypertensive heart disease, specified as benign, Hypertensive heart disease, not specified as malignant or benign, Hypertensive renal disease, Hypertensive renal disease, specified as malignant, Hypertensive renal disease, specified as benign, Hypertensive renal disease, not specified as malignant or benign, Hypertensive heart and renal disease, Hypertensive heart and renal disease, specified as malignant, Hypertensive heart and renal disease, specified as benign, Hypertensive heart and renal disease, not specified as malignant or benign, Secondary hypertension, Secondary hypertension, specified as malignant, Secondary hypertension, specified as benign, Secondary hypertension, not specified as malignant or benign |

**Figure 1.** Deep learning regression and segmentation model architectures

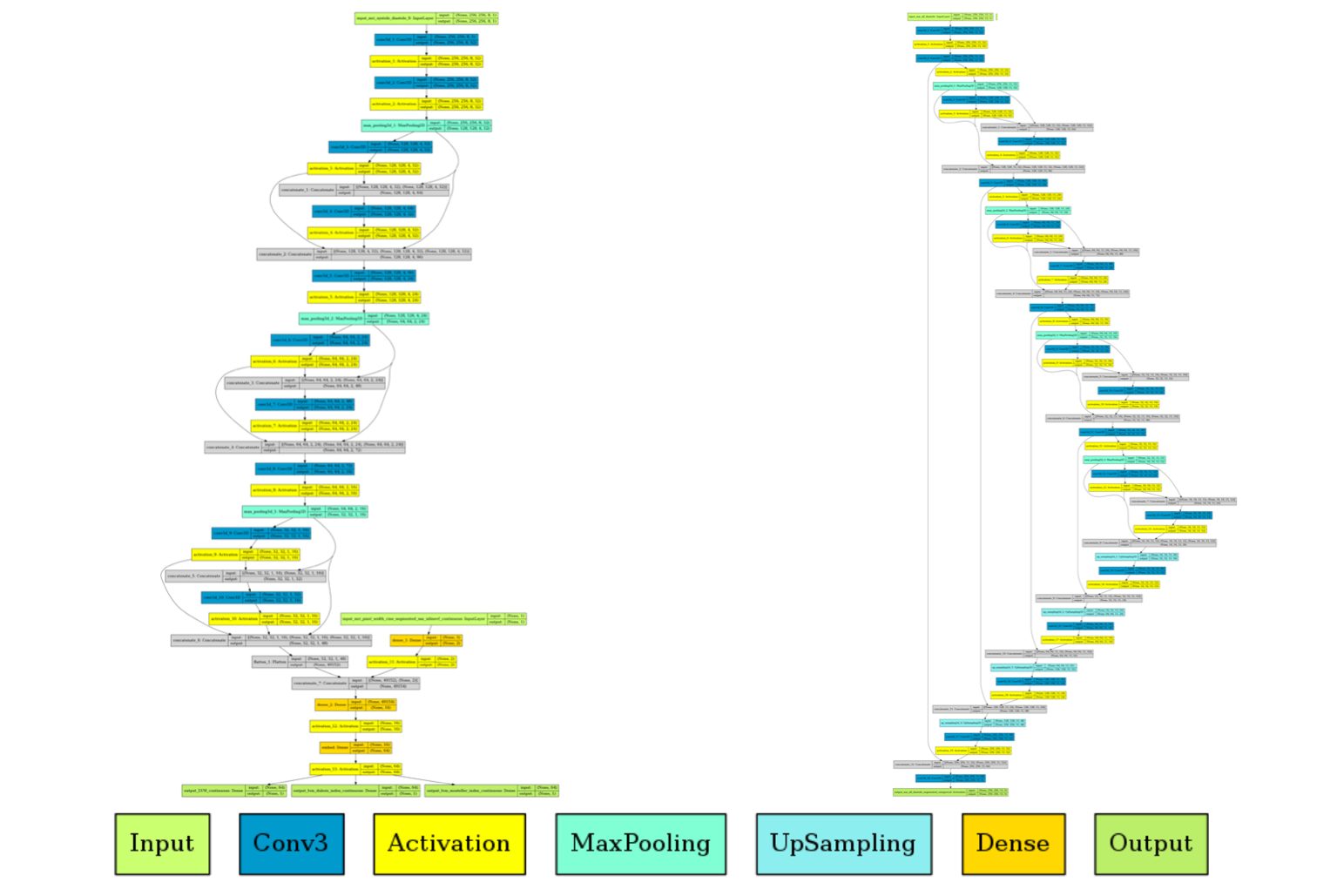

Depicted are overviews of deep learning model architecture. The left panel shows regression, with pixel size input and LVM as well as LVM indexed output. Right shows U-Net semantic segmentation architecture.

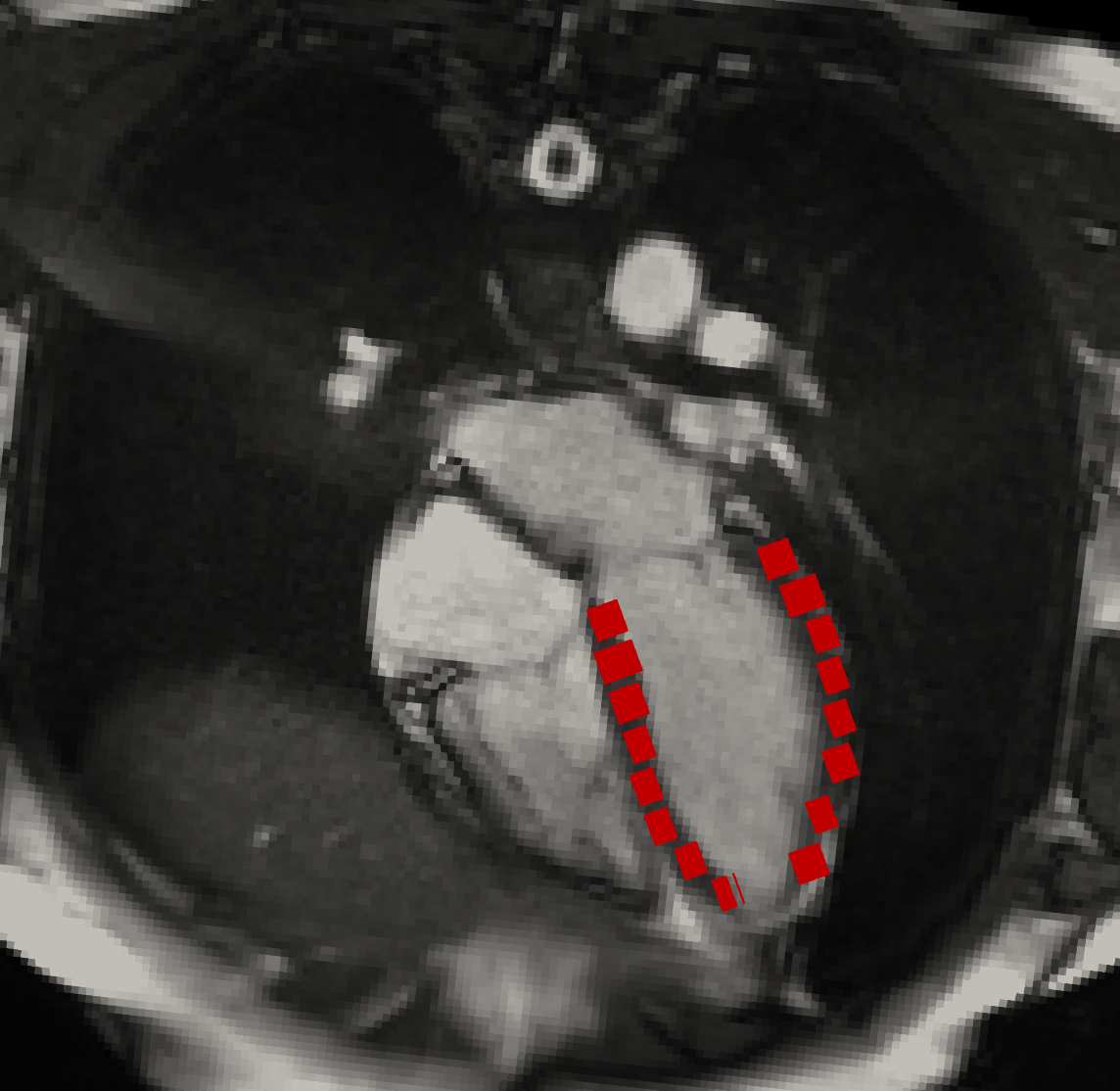
**Figure 2.** Determination of short-axis CMR slice thickness

Depicted is a representative rendering from one of the visual inspections performed to assess the thickness of short-axis CMR slices from DICOM metadata. InlineVF short-axis segmentations of the LV myocardium (rendered in red) are shown projected onto the 4-chamber long-axis CMR (rendered in grayscale) after a procedure of co-alignment performed via custom routines, which are now part of the open-source Machine Learning for Health (ML4H)^1^ package. More specifically, we first used image metadata from the standard *Image Position (Patient)*, *Image Orientation (Patient)*, and *Slice Thickness* DICOM tags to co-rotate the 4-chamber and short-axis views into the same reference system. Then, we sliced the short-axis segmentations along the plane of the 4-chamber CMR to allow for a visual assessment of the segmentation accuracy. As expected, the inlineVF segmentations match well the overall LV myocardium anatomy, as visible from the co-aligned long-axis 4-chamber CMR. However, small gaps are consistently interposed between adjacent segmentations, denoting how the distance between short-axis slices can be larger than the *Slice Thickness* value in the DICOM files. After inspecting the metadata of over 4,000 cases, we concluded that the distance between adjacent short-axis slices is equal to 10 mm in all but a handful of cases (24, i.e., < 0.6%), while the *Slice Thickness* metadata equaled 8 mm in all cases. Hence, to compensate for the 2-mm thickness gaps in our LV mass computations, we considered the short-axis CMR slices to have an out-of-plane size of 10 mm.

**Figure 3.** Saliency map of left ventricular mass semantic segmentation model

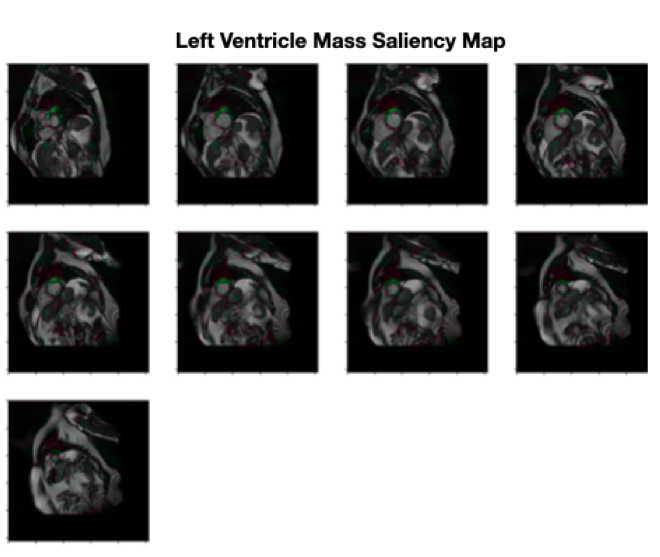

Depicted are saliency maps generated by the deep learning segmentation model. Areas highlighted in color depicts regions of the CMR appropriately recognized by the model as regions of left ventricular myocardium.

**Figure 4.** Correlation between manually labeled left ventricular mass and estimated LV mass using deep learning segmentation prior to adjustment

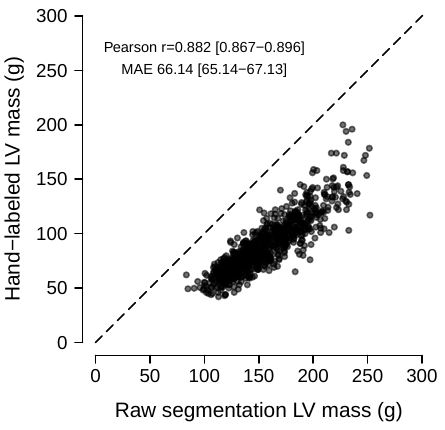

Depicted is the correlation between manually labeled LV mass (y-axis) and CMR-derived LV mass using deep learning segmentation prior to adjustment by mean centering in the test set independent of model training. Results depict a linear relationship between segmentation-based LV mass and manually labeled LV mass, but with bias due to systematic overestimation.

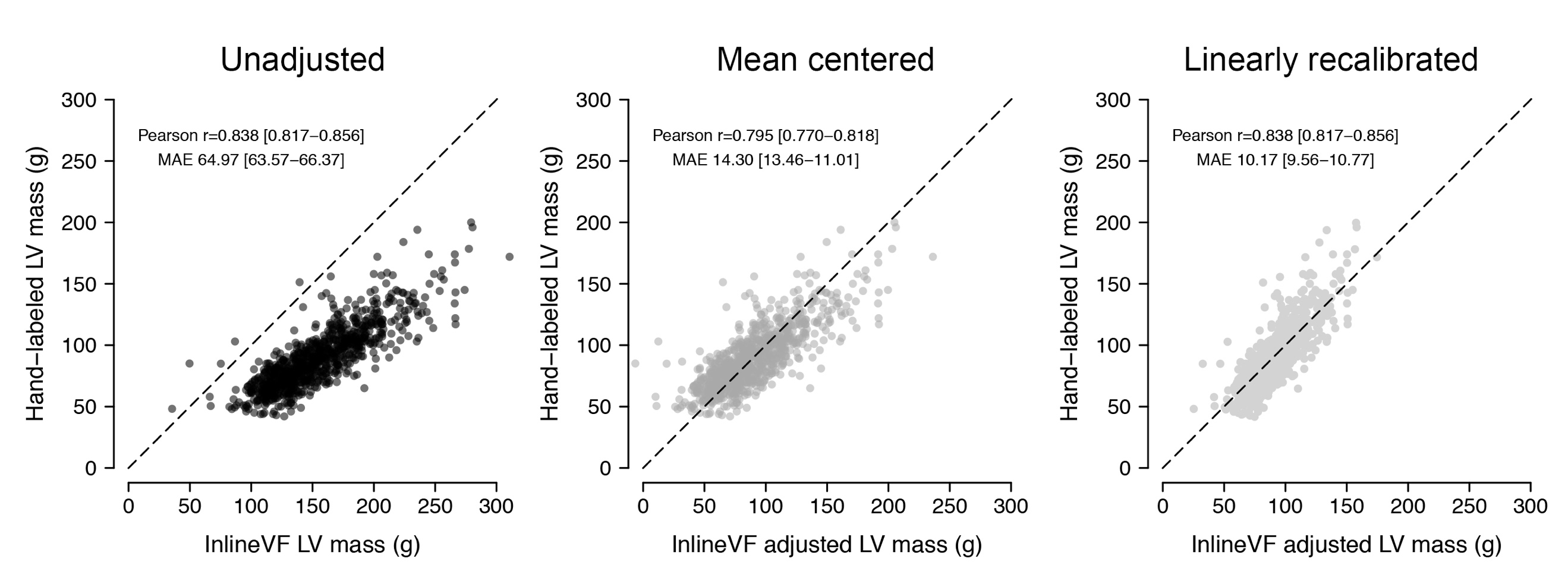
**Figure 5.** Correlation between manually labeled left ventricular mass and estimated LV mass using inlineVF with and without adjustment

Depicted are plots illustrating the correlation between manually labeled LV mass (y-axis) and CMR-derived LV mass using inlineVF with no adjustment (left panel), with centering upon the observed sex-stratified mean manually labeled LV mass (middle panel), and after full linear adjustment to manually labeled LV mass (right panel). Results are shown among individuals within the test set independent of model training (to allow direct comparison with the deep learning models).

**Methods.** Deep learning model training and evaluation

*Generation of pixel masks from segmentation contours*

InlineVF contours require processing to generate to pixel masks. The inner and outer contours of the myocardium may not form two distinct connected components, as they may intersect zero or more times, leading to ambiguity in the pixel mask label. To obtain a pixel mask from the contours we use the morphological closing operator with two circular kernels representing the minimum extent of the myocardium and maximum size of the blood pool (**Methods Figure 1**).

**Methods Figure 1**. Pixel masking using inlineVF contours

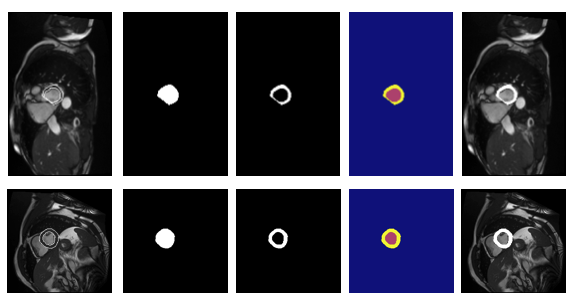

From left to right are depicted the polygons from InlineVF, morphological closing with maximum blood pool, morphological closing with minimum myocardium thickness disc, the summation of the two binary masks which is the class index specifying background = 0, ventricle = 1, and myocardium = 2, and lastly the myocardium label superimposed on the original MRI.

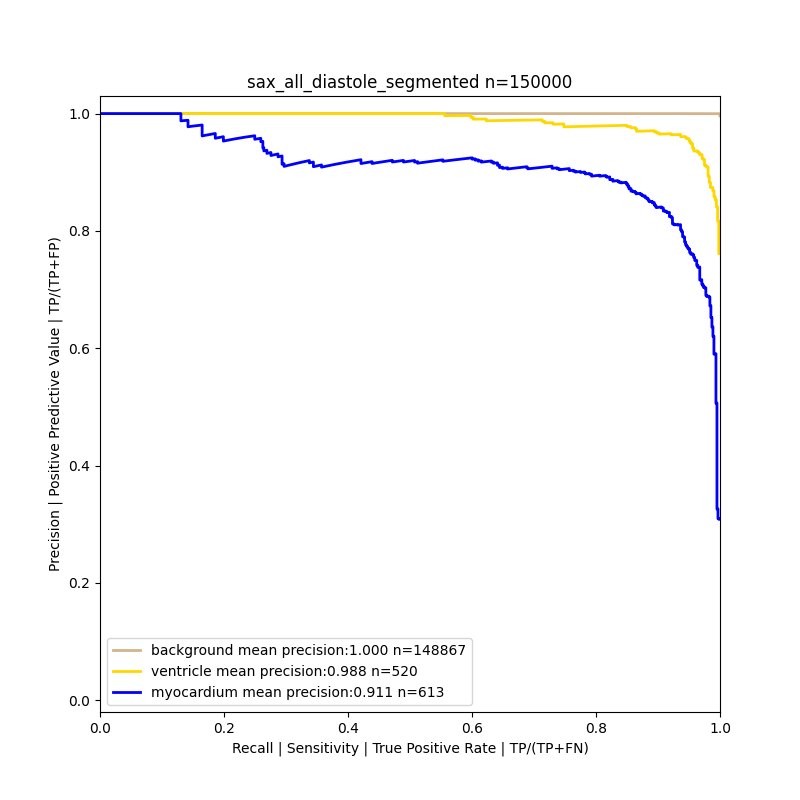
**Methods Figure 2**. Precision-recall curve of pixel identification

This curve plots the precision (y-axis) versus recall (x-axis) for identification of background (brown), ventricular blood pool (yellow), and myocardium (blue), evaluated within 150,000 voxels randomly selected from 5 MRIs in the test set. The class imbalance reflects smaller volume covered by the ventricle and myocardium.

*Model learning curves*

Training regression models on approximately 4,000 distinct manually labeled LVM values overfits in hours, demonstrated by the orange curve tracking validation set loss vs the blue which shows training set loss. On the other hand, training segmentation models on ~33K MRIs with pixel masks (i.e. billions of distinct values) even after days of training the model continues to improve slightly (**Methods Figure 3**).

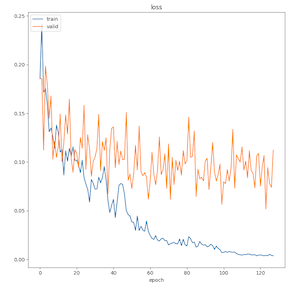
**Methods Figure 3.** Learning curves for regression and segmentation models

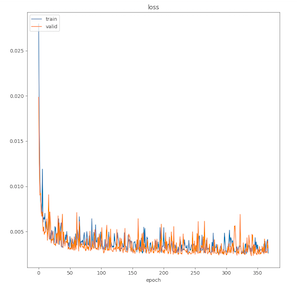

Depicted are learning curves plotting training and validation loss over epoch of training for the regression model (left) versus the segmentation model (right). Training loss is shown in blue and validation loss is shown in orange.

*Model calibration*

**Methods Figure 4**. Model calibration

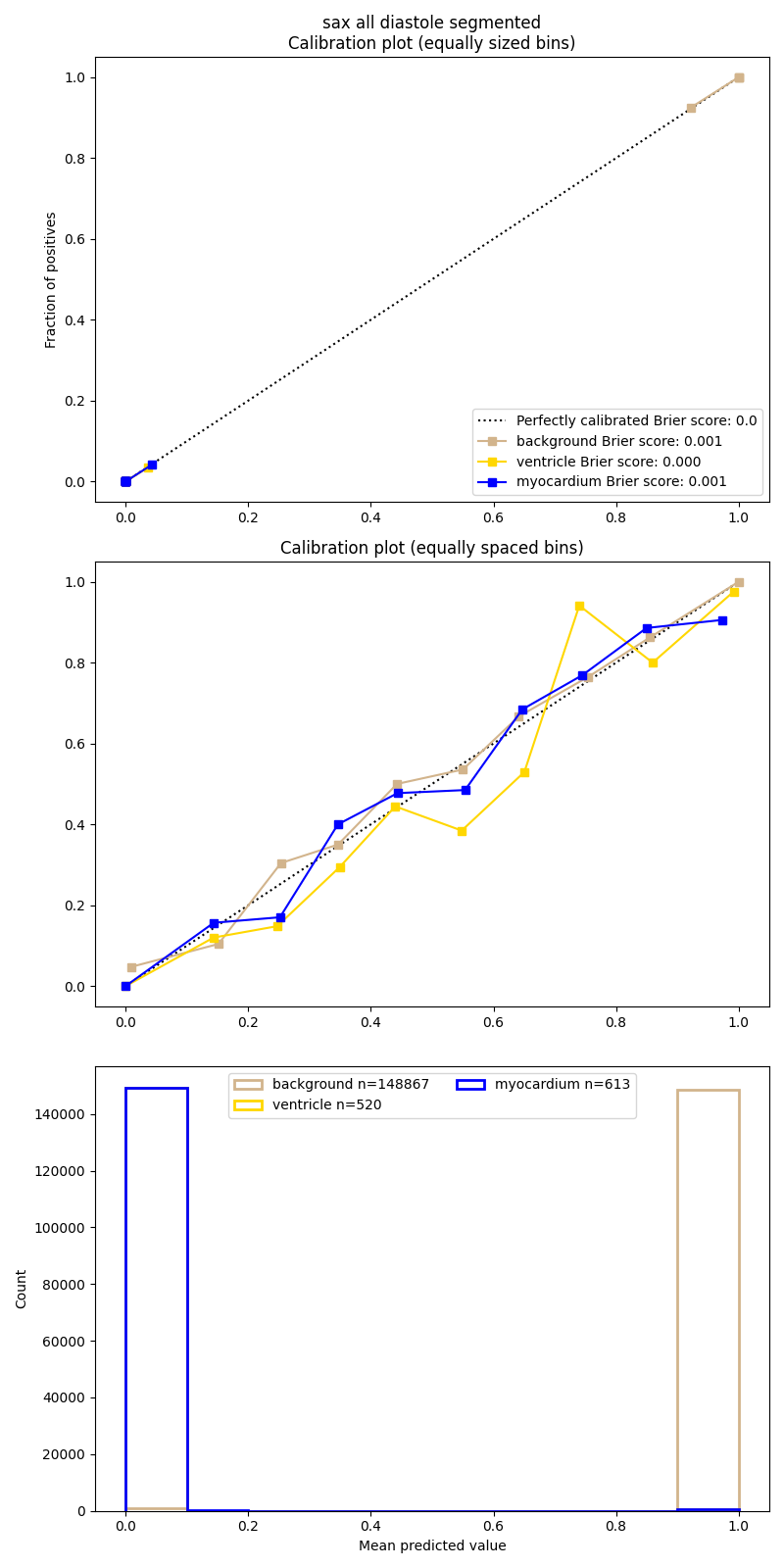

Plots depict calibration between model predictions of pixel identity (background – brown, ventricular blood pool – yellow, myocardium – blue) and true identity, calculated across voxel deciles (quantized into evenly sized bins in top plot vs quantized into evenly spaced bins in middle plot). The bottom panel shows the distribution of the model’s per-class predicted probabilities. While well-calibrated the model predictions are extremely bimodal, as is common with cross-entropy trained models.

**Supplemental References**

1.	ML4CVD Group. Machine Learning for Health (ML4H). https://github.com/broadinstitute/ml. *GitHub*. 2020;

2.	Petersen SE, Aung N, Sanghvi MM, Zemrak F, Fung K, Paiva JM, Francis JM, Khanji MY, Lukaschuk E, Lee AM, Carapella V, Kim YJ, Leeson P, Piechnik SK, Neubauer S. Reference ranges for cardiac structure and function using cardiovascular magnetic resonance (CMR) in Caucasians from the UK Biobank population cohort. *J Cardiovasc Magn Reson*. 2017;19:18.

3.	Loh P-R, Kichaev G, Gazal S, Schoech AP, Price AL. Mixed-model association for biobank-scale datasets. *Nat Genet*. 2018;50:906–908.

4.	Schizophrenia Working Group of the Psychiatric Genomics Consortium, Bulik-Sullivan BK, Loh P-R, Finucane HK, Ripke S, Yang J, Patterson N, Daly MJ, Price AL, Neale BM. LD Score regression distinguishes confounding from polygenicity in genome-wide association studies. *Nat Genet*. 2015;47:291–295.
